# The Impact of Intermittent Hypoxemia on Type 1 Retinopathy of Prematurity in Preterm Infants

**DOI:** 10.1101/2023.09.25.23295922

**Authors:** Samaneh Rabienia Haratbar, Li Chen, Qiang Cheng, Dara Singh, Faraneh Fathi, Mehrana Mohtasebi, Xuhui Liu, Abhijit Patwardhan, Prasad Bhandary, Henrietta S. Bada, Guoqiang Yu, Elie G. Abu Jawdeh

**Affiliations:** Department of Biomedical Engineering, University of Kentucky; Biostatistics and Bioinformatics Shared Resource Facility, Markey Cancer Center, University of Kentucky; Institute for Biomedical Informatics, Department of Internal Medicine and Department of Computer Science; Division of Neonatology, Department of Pediatrics, University of Kentucky

**Author notes:** **Corresponding Authors:** Elie G. Abu Jawdeh, MD, PhD Address: 138 Leader Ave, 010A, Lexington, KY, USA 40536-0679, Guoqiang Yu, PhD Address: HKRB #117, 760 Press Avenue, Lexington, KY, USA 40536-0679.

## Abstract

**Background:** Intermittent hypoxemia (IH) may influence retinopathy of prematurity (ROP) development in preterm infants, however, previous studies had mixed results. This study aims to assess the influence and evaluate the predictive ability of IH measures on Type 1 ROP, a stage beyond which ROP treatment is indicated.

**Methods:** IH was quantified by continuously monitoring oxygen saturation (SpO_2_) using high-resolution pulse oximeters during the first 10 weeks of life. Statistical analyses assessed the relationship and predictive ability of weekly and cumulative IH variables for Type 1 ROP development.

**Results:** Univariate analyses suggested that IH measures are greater in infants with Type 1 ROP and are predictive of Type 1 ROP development. Multivariable logistic regression analyses revealed that cumulative IH of longer duration during certain postnatal periods are associated with Type 1 ROP development after adjusting for gestational age (GA) or birth weight (BW). Although area under the curve (AUC) analyses revealed added predictivity of cumulative IH variables above GA or BW, these increments in AUC were not statistically significant.

**Conclusions:** The duration of IH events was associated with Type 1 ROP development. Interventions for reducing the duration of IH events may potentially improve ROP outcomes.

**Impact:**

- This study investigates the impact of IH on the development of Type 1 ROP in preterm infants.
- Univariate analyses revealed that IH measures are greater in infants with Type 1 ROP and are predictive of Type 1 ROP development.
- Multivariable logistic regression analyses revealed that cumulative IH events of longer duration are associated with Type 1 ROP development after adjusting for GA or BW.
- Interventions for reducing the duration of IH events during critical postnatal periods may potentially improve ROP outcomes.

## 1. Introduction

Retinopathy of prematurity (ROP) remains a leading cause of visual impairment in children, despite early detection and treatment advancements^1,2^. Multiple risk factors such as prematurity, low gestational age (GA), low birth weight (BW), and need for mechanical ventilation, are associated with the development of ROP^2–6^. Most of these factors may not be modifiable by the managing neonatology team. Identification of a modifiable risk factor has significant clinical relevance as it has the potential to improve outcomes for ROP.

Intermittent hypoxemia (IH) is generally defined as episodic drops in hemoglobin oxygen saturation (SpO_2_). Brief episodes of oxygen desaturation may seem clinically insignificant, but these events occurring up to hundreds of times per day, may have a cumulative effect on neonatal morbidities, including ROP in preterm infants^7^. Two research groups have previously reported a positive relationship between the IH and ROP. Di Fiore et al. showed extremely preterm infants with severe ROP requiring laser therapy had higher IH frequency compared to infants who did not require surgery^8,9^. Poets et al. in a post hoc analyses of the Canadian Oxygen Trial showed infants with severe ROP significantly spent more time in hypoxemia compared to infants without severe ROP^10^. On the other hand, Fairchild et al. in a large single center study showed no relationship between IH measures and ROP in preterm infants^11^. Given the variation in literature, in this study, we readdress these relationships and assess the predictive ability of IH in the development of Type 1 ROP, a stage beyond which ROP treatment is indicated.

## 2. Methods

### 2.1 Study Design and Population

This study was approved by the University of Kentucky’s Institutional Review Board. Preterm infants between 23 0/7 to 30 6/7 weeks GA admitted to the neonatal intensive care unit (NICU) were included in the study. Infants were enrolled prospectively upon admission and consented for high resolution data collection from research pulse oximeters. Infants with major congenital malformations such as congenital heart disease were excluded as they may have other reasons for hypoxemia. Demographic information was retrospectively obtained by review of medical records. The staging of ROP was obtained from ophthalmology exam documentation.

### 2.2 Data Collection

Oxygen saturation data (SpO_2_) were collected from the bedside using high-resolution (2 second averaging time and a 1 Hz sampling rate) pulse oximeters (Radical 7; Masimo, Irvine, CA, USA) to continuously monitor patients through 10 weeks postnatal age^12–14^. Pulse oximeters were equipped with serial data recorders (Acumen Instruments, Ames, IA, USA) for continuous data collection^12–14^. Novel programs were created through MATLAB (Natick, MA, USA) to filter and analyze the alterations in IH measures^12,14,15^. Data with artifacts or signal loss were excluded; only SpO_2_ data with good signals were included in the analyses. IH outcomes were calculated as a drop in SpO_2_ to less than 80%. The number and duration of IH events and percent time with SpO_2_ < 80% were quantified daily for each infant and reported cumulatively over the study period. Type 1 ROP is defined as a stage beyond which ROP treatment is required, including zone 1 ROP with plus disease, zone 1 and stage 3 ROP without plus disease, and zone 2 and stage 2 or stage 3 ROP with plus disease^16^.

### 2.3 Data Process and Statistical Analysis

Daily IH data were processed for weekly and cumulative analyses. Results between the Type 1 ROP and non-Type 1 ROP groups were compared week by week and cumulative multiple weeks. Two cumulative processes were conducted including process #1, in which data were calculated by adding each week to the sum of its predecessors (starting from week 10) and process #2, in which data were calculated by adding each week to the sum of its successors (starting from week 1).

Statistical analysis of all measures was performed using SPSS program version 28.0 (SPSS, Inc., Chicago, IL, USA). Results were considered statistically significant with p < 0.05. The Mann-Whitney U tests were used to compare the values of IH variables between Type 1 ROP and non-Type 1 ROP groups. The multivariable logistic regression analyses were performed to assess the association between IH variables and the development and progression of ROP in these infants after adjusting for GA or BW. The receiver operating characteristic (ROC) curve and the area under the ROC curve (AUC) were used for evaluating the predictive ability of IH variables in the prediction of Type 1 ROP. The AUC of combining each IH variable and GA or BW, obtained from the multivariable logistic regression model, was compared to the AUC of GA or BW to evaluate whether this IH variable has added predictive ability above GA or BW. The ROC curve plots sensitivity (probability of a true positive) against one minus specificity (probability of a false positive) of a risk factor for all possible threshold values^17^. The AUC quantifies the overall predictive ability of a risk factor.

## 3. Results

The study population included 210 infants: 30 with Type 1 ROP and 180 non-Type 1 ROP. Table 1 shows the subject demographics and baseline characteristics. Data are presented as mean ± standard error (SEM) and n (percent) for continuous and categorical variables, respectively. The infants with Type 1 ROP were of lower GA (24.4 ± 1.3 weeks versus 27.0 ± 1.7 weeks, p < 0.001) and lower BW (694.0 ± 162.9 grams versus 1003.0 ± 269.6 grams, p < 0.001). There were females (49%) and males (51%) in the study population.

**Table 1:** Demographics and baseline characteristics.

|  | Type 1 ROP<br>(n = 30) | Non-Type 1 ROP<br>(n = 180) | p value |
| --- | --- | --- | --- |
| Gestational Age (weeks)<sup>a</sup> | 24.4 ± 1.3 | 27.0 ± 1.7 | **< 0.001** |
| Birth Weight (grams)<sup>a</sup> | 694.0 ± 162.9 | 1003.0 ± 269.6 | **< 0.001** |
| Race/Ethnicity |  |  |  |
| Non-Hispanic/White race, n (%) | 30 (100%) | 152 (84%) | 0.5 |
| Hispanic/White race, n (%) | 0 | 5 (3%) |  |
| Non-Hispanic/Asian race, n (%) | 0 | 2 (1%) |  |
| Non-Hispanic/Black race, n (%) | 0 | 21 (12%) |  |
| Male sex, n (%) | 17 (57%) | 84 (48%) | 0.3 |
| Multiple births, n (%) | 11 (17%) | 53 (30%) | 0.7 |
| Prenatal steroids, n (%) | 29 (97%) | 166 (92%) | 0.5 |
| Cesarean section, n (%) | 19 (64%) | 117 (65%) | 0.8 |

Note: The p values for continuous characteristics were calculated based on the Mann-Whitney U tests. The p values for categorical characteristics were calculated based on Chi-square tests or Fisher's exact tests, as appropriate. Bold numbers indicate significant p-values ( $p < 0.05$ ) comparing between the two groups of Type 1 ROP and non-Type 1 ROP.
<sup>a</sup> Mean ± SEM

### 3.1 Weekly Process

Figure 1 shows weekly mean values of IH variables for Type 1 ROP and non-Type 1 ROP and AUC values of IH variables. The results shown in Figure 1a-1c indicate that IH variables (i.e., the percent time with hypoxemia, duration of IH events, and number of IH events) had significant differences between Type 1 ROP and non-Type1 ROP groups during specific weeks. The percent time with hypoxemia demonstrated significant differences in week 1 and from week 4 to week 10 (p < 0.05); the duration of IH events showed significant differences from week 2 to week 9 (p < 0.05); and the number of IH events showed significant differences in week 1 and week 9 (p < 0.05). The AUC results shown in Figure 1d-1f reveal that those IH variables during aforementioned weeks also demonstrated significant predictive ability, with the duration of IH events having the highest AUC of 0.81 at peak IH in week 6 (Figure 1e).

**Figure 1:**
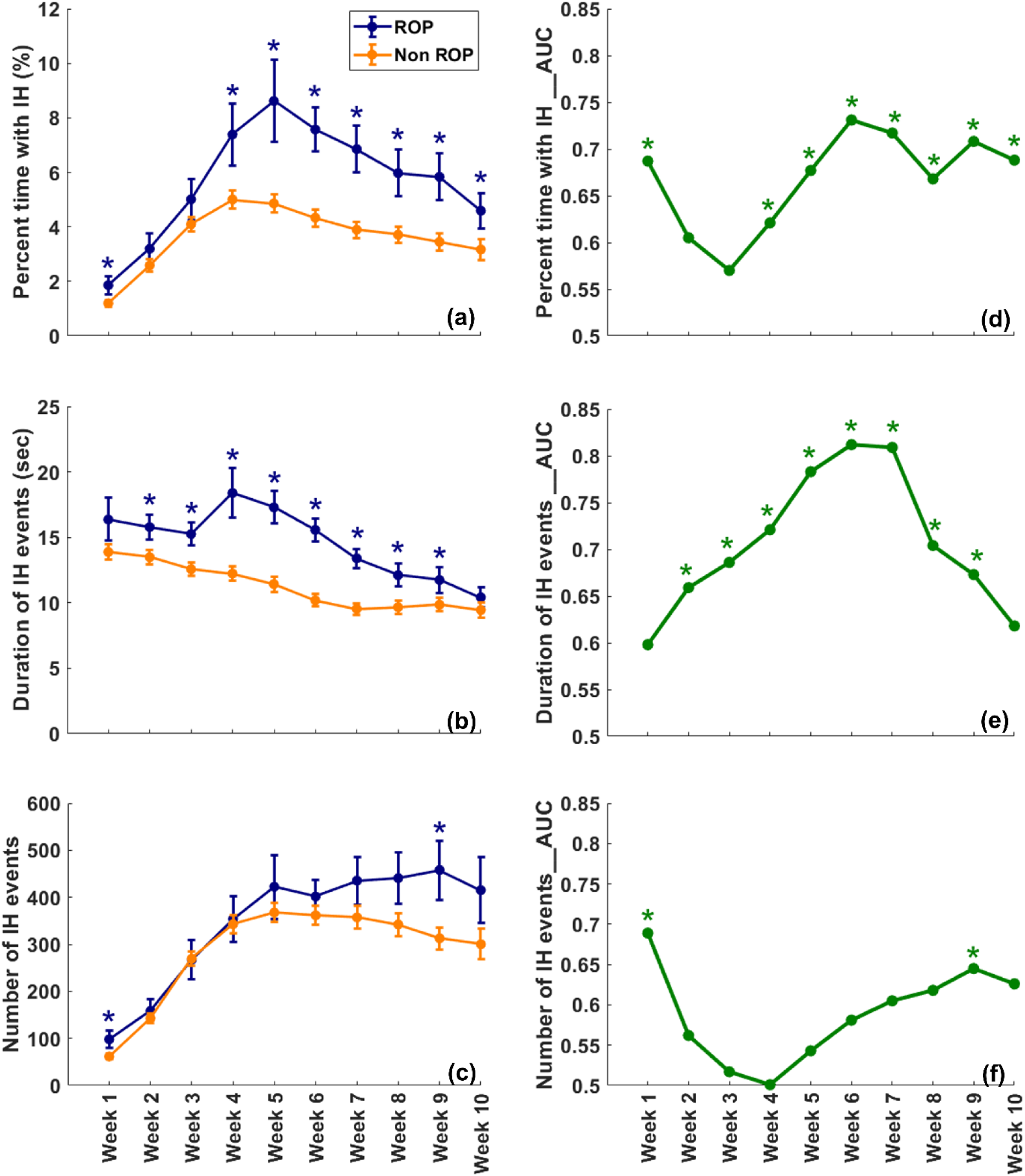
Weekly data analysis results. (a)-(c): Weekly comparison results (mean value ± SEM) between the Type1 ROP and non-Type1 ROP groups in IH variables, including the percent time with hypoxemia, duration of IH events, and number of IH events, respectively. * Indicates p < 0.05 for comparing the IH values between the two groups based on the Mann-Whitney U test. (d)-(f) Weekly AUC results in IH variables, including the percent time with hypoxemia, duration of IH events, and number of IH events, respectively. * Indicates p < 0.05 for assessing whether a single variable is predictive (i.e., AUC > 0.5) based on a test which is equivalent to the Mann-Whitney U test.

Multivariable logistic regression analysis was then performed using each of the IH variables and each of the risk factors (GA and BW). The objective was to determine whether each of IH variables is associated with ROP status while considering the influence of GA or BW adjustments. Table 2 shows adjusted p values based on multivariable logistic regression models. Results indicate that weekly IH variables were not significantly associated with ROP status after adjusting for GA except for the number of IH events at peak IH (week 4, p = 0.045). Similarly, weekly IH variables were not significantly associated with ROP status after adjusting for BW. The AUC analyses reveal that, overall, weekly IH variables had no considerable added predictive ability above GA and BW, where the AUC increases ranged from 0 - 0.01.

**Table 2:**
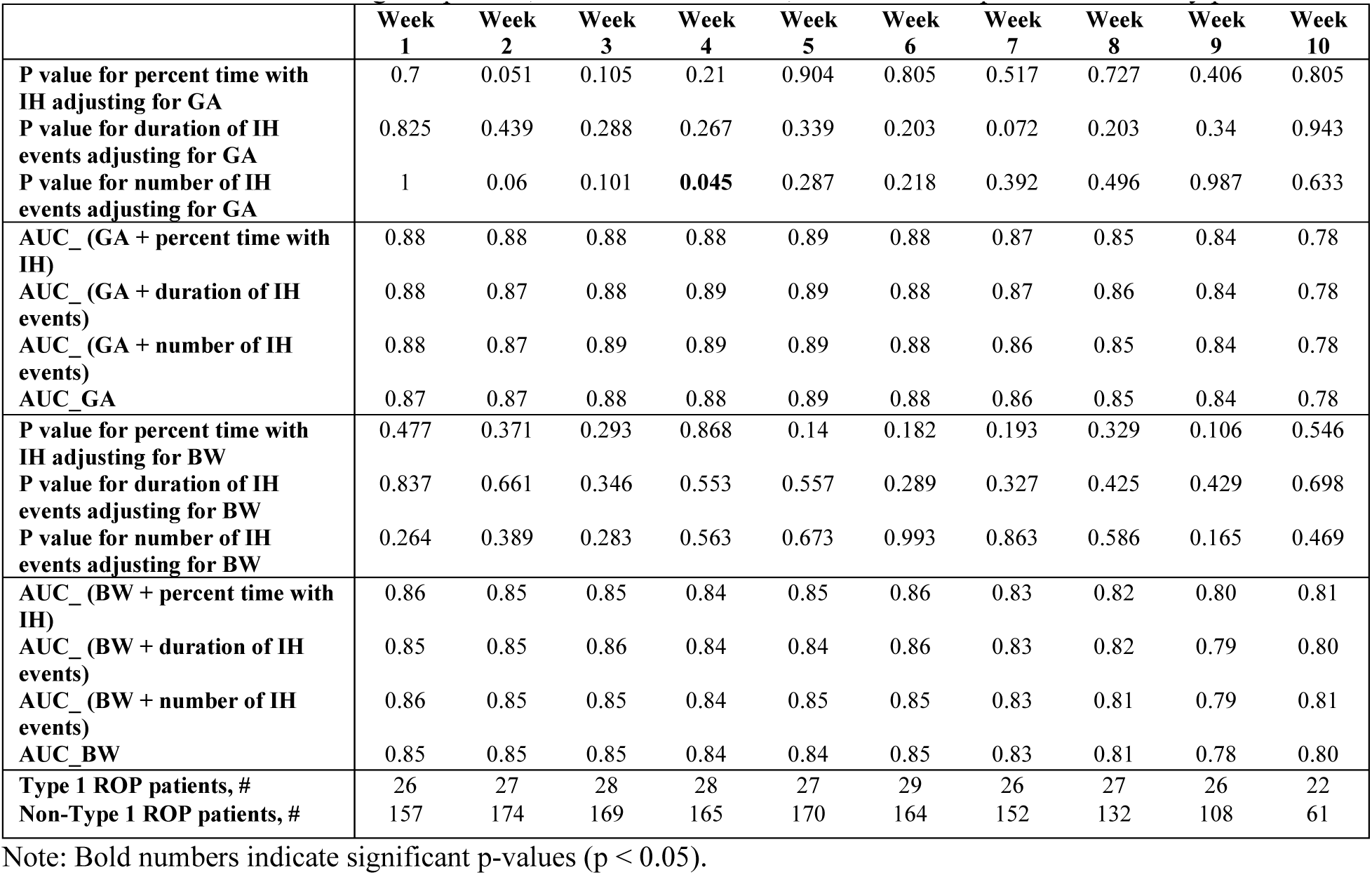
Multivariable logistic p value, AUC of IH variables, and number of patients for weekly process.

### 3.2 Cumulative Process

Figure 2 shows the mean values of IH variables for Type 1 ROP and non-Type 1 ROP and AUC values of IH variables for the cumulative process #1. The results shown in Figure 2a-2c indicate that the percent time with hypoxemia demonstrated significant differences between the two groups of Type 1 ROP and non-Type 1 ROP from cumulative week 5-10 to week 10 (p < 0.05) and the duration of IH events demonstrated significant differences from cumulative week 1-10 to week 9-10 (p < 0.05). The AUC results shown in Figure 2d-2f reveal that the percent time with hypoxemia and the duration of IH events during the aforementioned cumulative weeks also demonstrated significant predictive ability, with the duration of IH events having the highest AUC of 0.82 in cumulative week 5-10 (Figure 2e). The number of IH events had no significant difference between the two groups of Type 1 ROP and non-Type 1 ROP and no significant predictive ability for the development of Type 1 ROP.

**Figure 2:**
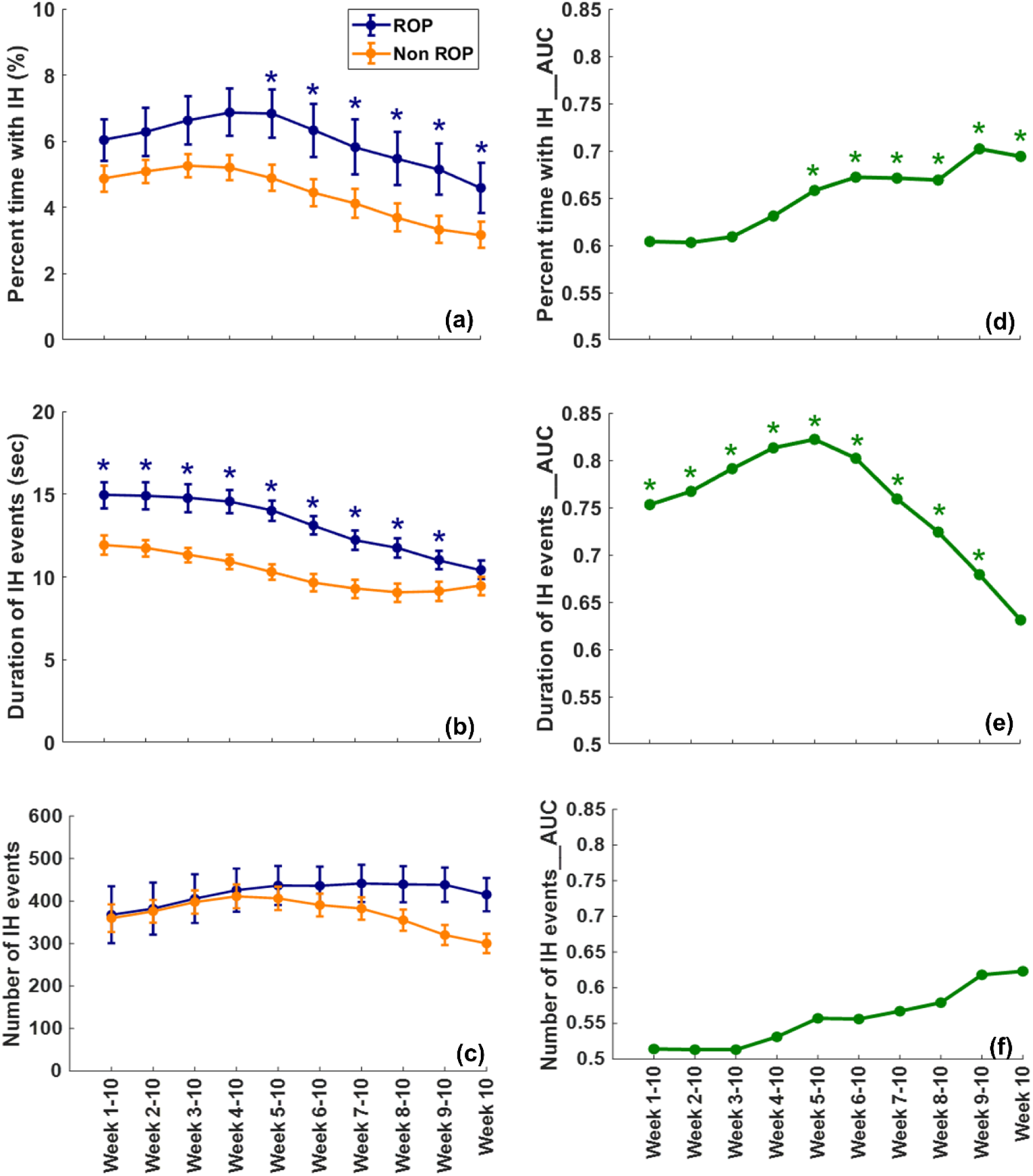
Cumulative process #1 results. (a)-(c): Cumulative comparison results (mean value ± SEM) between the ROP and non-Type1 ROP groups in IH variables, including the percent time with hypoxemia, duration of IH events, and number of IH events, respectively. * Indicates p < 0.05 for comparing the IH values between the two groups based on the Mann-Whitney U test. (d)-(f) Cumulative AUC results in IH variables, including the percent time with hypoxemia, duration of IH events, and number of IH events, respectively. * Indicates p < 0.05 for assessing whether a single IH variable is predictive (i.e., AUC > 0.5) based on a test which is equivalent to the Mann-Whitney U test.

Table 3 shows the adjusted p values based on multivariable logistic regression models for cumulative process #1. The results indicate that the cumulative duration of IH events was significantly associated with ROP status after adjusting for GA from cumulative week 3-10 to cumulative week 7-10 (p < 0.05). Similarly, the cumulative duration of IH events was significantly associated with ROP status after adjusting for BW from cumulative week 3-10 to cumulative week 8-10 (p < 0.05). The percent time with hypoxemia and the number of IH events did not show significant association with ROP status after adjusting for GA or BW.

**Table 3:** Multivariable logistic p value, AUC of IH variables and number of patients for cumulative process #1.

|  | Week<br>1-10 | Week<br>2-10 | Week<br>3-10 | Week<br>4-10 | Week<br>5-10 | Week<br>6-10 | Week<br>7-10 | Week<br>8-10 | Week<br>9-10 | Week<br>10 |
| --- | --- | --- | --- | --- | --- | --- | --- | --- | --- | --- |
| P value for percent time with IH<br>adjusting for GA | 0.624 | 0.638 | 0.754 | 0.948 | 0.79 | 0.739 | 0.774 | 0.666 | 0.471 | 0.829 |
| P value for duration of IH events<br>adjusting for GA | 0.072 | 0.067 | **0.032** | **0.019** | **0.016** | **0.014** | **0.03** | 0.052 | 0.425 | 0.969 |
| P value for number of IH events<br>adjusting for GA | 0.123 | 0.127 | 0.126 | 0.156 | 0.23 | 0.33 | 0.492 | 0.74 | 0.837 | 0.7 |
| AUC_ (GA + percent time with<br>IH) | 0.79 | 0.79 | 0.79 | 0.79 | 0.79 | 0.80 | 0.79 | 0.79 | 0.79 | 0.79 |
| AUC_ (GA + duration of IH<br>events) | 0.81 | 0.81 | 0.81 | 0.82 | 0.83 | 0.84 | 0.82 | 0.80 | 0.80 | 0.78 |
| AUC_ (GA + number of IH<br>events) | 0.79 | 0.78 | 0.78 | 0.78 | 0.78 | 0.78 | 0.79 | 0.79 | 0.79 | 0.78 |
| AUC_GA | 0.79 | 0.79 | 0.79 | 0.79 | 0.79 | 0.79 | 0.79 | 0.79 | 0.79 | 0.79 |
| P value for percent time with IH<br>adjusting for BW | 0.634 | 0.653 | 0.556 | 0.367 | 0.2 | 0.213 | 0.231 | 0.158 | 0.16 | 0.526 |
| P value for duration of IH events<br>adjusting for BW | 0.109 | 0.095 | **0.047** | **0.027** | **0.015** | **0.009** | **0.016** | **0.023** | 0.25 | 0.56 |
| P value for number of IH events<br>adjusting for BW | 0.756 | 0.711 | 0.73 | 0.875 | 0.915 | 0.81 | 0.618 | 0.413 | 0.255 | 0.512 |
| AUC_ (BW + percent time with<br>IH) | 0.79 | 0.79 | 0.79 | 0.80 | 0.80 | 0.80 | 0.80 | 0.80 | 0.81 | 0.81 |
| AUC_ (BW + duration of IH<br>events) | 0.80 | 0.80 | 0.81 | 0.81 | 0.83 | 0.83 | 0.83 | 0.82 | 0.80 | 0.80 |
| AUC_ (BW + number of IH<br>events) | 0.78 | 0.78 | 0.78 | 0.78 | 0.79 | 0.79 | 0.79 | 0.79 | 0.80 | 0.81 |
| AUC_BW | 0.79 | 0.79 | 0.79 | 0.79 | 0.79 | 0.79 | 0.79 | 0.79 | 0.79 | 0.79 |
| Type 1 ROP patients, # | 24 | 24 | 24 | 24 | 24 | 24 | 24 | 24 | 24 | 24 |
| Non-Type 1 ROP patients, # | 62 | 62 | 62 | 62 | 62 | 62 | 62 | 62 | 62 | 62 |

Note: Bold numbers indicate significant p values ( $p < 0.05$ ).

The AUC analyses reveal that the duration of IH event had AUC increases ≥ 0.03 above GA from cumulative week 4-10 to cumulative week 7-10. The duration of IH event had AUC increases ≥ 0.03 above BW from cumulative week 5-10 to cumulative week 8-10. However, these AUC increases were not statistically significant (p > 0.05; data not shown). The percent time with hypoxemia and the number of IH events had no considerable added predictive ability above GA or BW, where the AUC increases ranged from 0 - 0.02.

The cumulative process #2 generated similar results (Supplementary Figure 1). The results indicate that IH variables had significant difference between the two groups of Type 1 ROP and non-Type 1 ROP. The percent time with hypoxemia had significant differences in week 1, cumulative week 1-2, and from cumulative week 1-5 to cumulative week 1-9 (p < 0.05); the duration of IH events had significant differences from cumulative week 1-2 to cumulative week 1-10 (p < 0.05); and the number of IH events had a significant difference in week 1 (p <0.05). Those IH variables also showed significant predictive ability during the aforementioned cumulative weeks, with the duration of IH events having the highest AUC of 0.77 in cumulative week 1-7 (Supplementary Figure 1e).

Adjusted p values based on multivariable logistic regression models for cumulative process #2 (Supplementary Table 1) indicate that accumulating IH variables were not significantly associated with ROP status after adjusting for GA except for the percent time with hypoxemia in cumulative week 1-2. The AUC analyses reveal that IH measures in cumulative process #2 had no considerable added predictive ability above GA or BW, where the AUC increases ranged from 0 - 0.01.

## 4. Discussion

The present study describes positive relationships between IH measures and Type 1 ROP in preterm infants less than 31 weeks GA. Mainly, our results show the impact of the duration of IH events on the development of Type 1 ROP. This important finding is exciting as the extent of IH may be modifiable with prompt adjustment of oxygen supplementation needs in preterm infants, in addition to the optimization of methylxanthine therapy and respiratory support. Multiple studies have assessed factors for the risk of ROP in preterm infants, which however are mostly non-modifiable factors such as low GA, low BW, and the need for respiratory support.

Intermittent hypoxemia measures, mainly percent time in hypoxemia and duration of IH events, were associated with Type 1 ROP in both the weekly (Figure 1) and cumulative process (Figure 2). The duration of cumulative IH events in cumulative Process #1 remained significant after adjusting for GA and BW. That is, the longer IH events added up after 3 weeks of life and were associated with Type 1 ROP (Table 3). In contrast, results from cumulative Process #2 were not significantly associated, likely due to being remote from Type 1 ROP progression. We previously reported IH changes with postnatal age in this preterm infant population; there was low number of IH events in the first week of life, then an increase in IH events to a peak at 4 - 6 weeks of life followed by a plateau^7,8,18^. This peak in IH events may be the most clinically relevant, and intervening to decrease the duration of IH events during that period may improve outcomes.

In addition to the relationship between IH and Type 1 ROP, we assessed the predictive ability of IH measures on Type 1 ROP. We found that IH measures were predictive of Type 1 ROP (Figure 1 and Figure 2). There were also some considerable increases in AUC (≥ 0.03) above GA or BW for cumulative process #1 (Table 3), albeit not statistically significant, likely due to the small sample size. In the future, we plan to expand our cohort and consider machine learning techniques to evaluate IH patterns that predict Type 1 ROP. As previously mentioned, the duration of IH events may be a modifiable ROP risk factor and a mild to moderate increase in predictive ability may be clinically relevant.

We selected Type 1 ROP (also known as high-risk pre-threshold ROP) as our outcome of interest since this classification is the first indication for treatment regardless of whether the patient received therapy. Our outcome measure is different from previous studies that focused on either ROP therapy or severe ROP^8–11^. Our study also included all preterm infants less than 31 weeks, as this patient population is routinely screened for ROP and has a high IH burden^18^. In contrast, Di Fiore et al. and Poets et al. focused on extremely preterm infants less than 28 weeks GA. We utilized data acquired from high-resolution pulse oximeters set to 2 second averaging time and 1 Hz sampling rate. Poets et al.^10^ and Fairchild et al.^11^ utilized monitors with averaging time set to 16 seconds and 8 seconds, respectively. Smaller averaging times allow for more precisely quantifying IH measures including those of short and long durations^12,19^. Longer averaging times may underestimate the number of short IH events and overestimate IH of longer durations^12,19^. It is possible that these differences in patient population, outcomes measures, and pulse oximeter averaging times influenced some of the results among studies.

One limitation of this study is the data analysis of an existing database at a single center. The small sample size did not allow for extensive multivariable analyses and adjustments and hence could not specify a duration beyond which Type 1 ROP risk increases. Future studies with larger sample size at multiple centers can focus on identifying a pattern or threshold beyond which IH predicts ROP. This is important as IH duration may be a modifiable ROP risk factor with prompt or automated adjustment of oxygen supplementation^20^. Also, future studies can focus on earlier severity stages of ROP wherein early intervention for IH optimization may have a great impact on ROP development.

## 5. Conclusions

This study investigated the relationship between the IH variables and the development of Type 1 ROP in preterm infants. The findings suggest that the duration of IH events was significantly associated with Type 1 ROP development. The study also explored the predictive ability of IH measures for Type 1 ROP and found that IH measures were predictive. The added predictive ability of IH measures above GA or BW, although considerable, was not statistically significant likely due to the small sample size. Our study suggests that IH may increase the ROP risk, and interventions for reducing the duration of IH events during critical postnatal periods may potentially improve ROP outcomes. Our findings contribute to the understanding of the impact of IH duration on the development of Type 1 ROP, offering insights for potential interventions to improve visual outcomes in vulnerable preterm infants.

## Data Availability

All data produced in the present study are available upon reasonable request to the authors

## Data Availability Statement

The data that support the findings of this study are available from the corresponding author upon reasonable request.

## Acknowledgments

We thank the research coordinators and research staff for assisting with subject enrollment and data collection. Special thank you to Sara Butler, RN, CCCE, and Crystal Wilson, LPN.

## Author Contributions

E.G.A, G.Y., H.S.B, and S.R.H conceived the study. E.G.A conducted and supervised human experiments. E.G.A, and P.B acquired data. S.R.H, and Li C. analyzed and interpreted data. S.R.H, and L.C. performed statistical analysis. E.G.A, P.B., and H.S.B provided resources for human experiments. S.R.H. drafted the manuscript. D.S., F.F., M.M., X.L., G.Y., L.C., E.G.A, H.S.B., Q.C., A.P., P.B., and S.R.H reviewed and edited the manuscript. All authors gave final approval of the version to be published.

## Consent Statement

Under a protocol approved by the Institutional Review Board of the University of Kentucky, informed consent was obtained by the research team from each subject’s parent.

---

**Supplementary Table 1:**
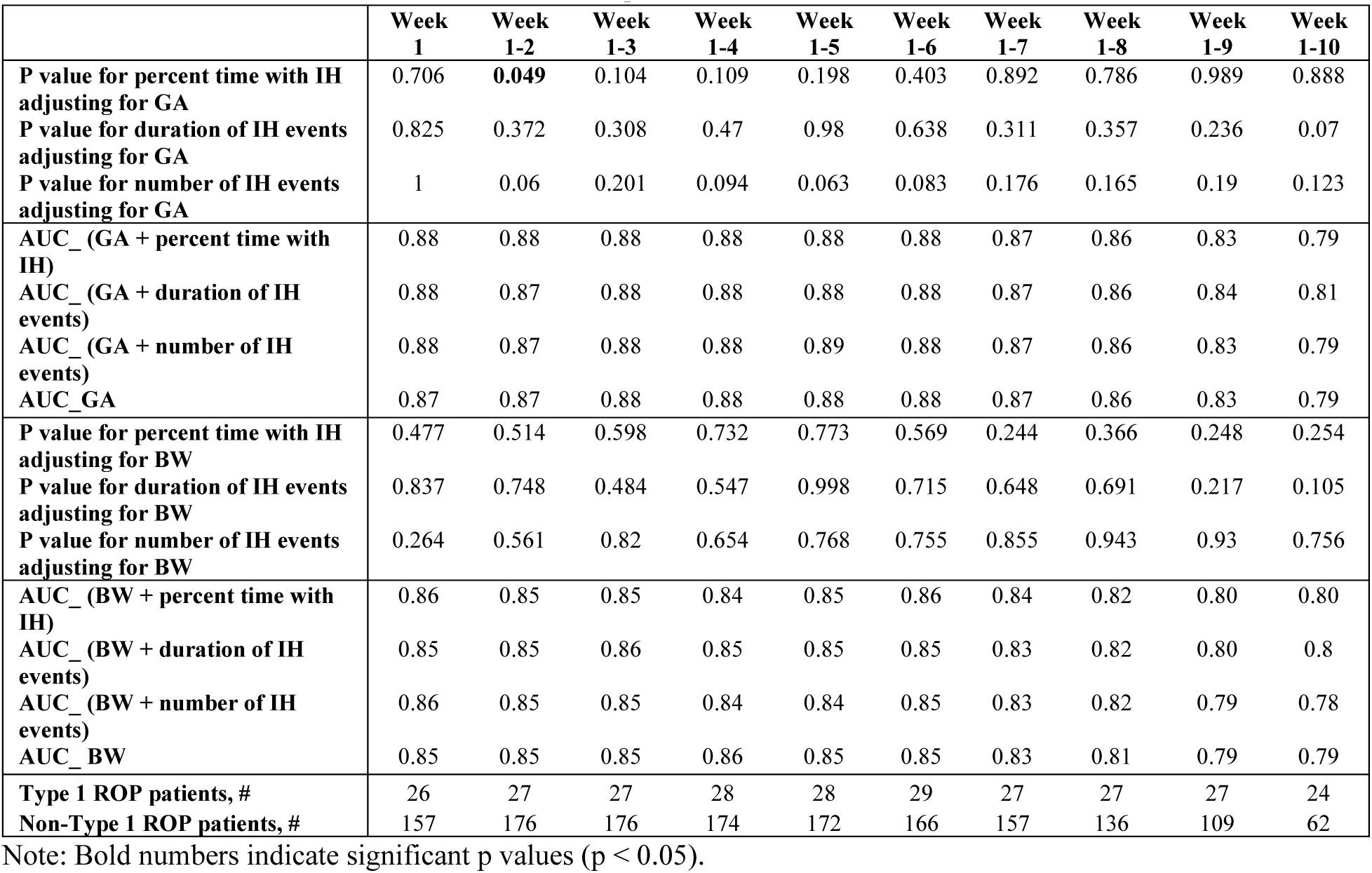
Multivariable logistic p value, AUC of IH variables and number of patients for cumulative process #2.

**Supplementary Figure 1:**
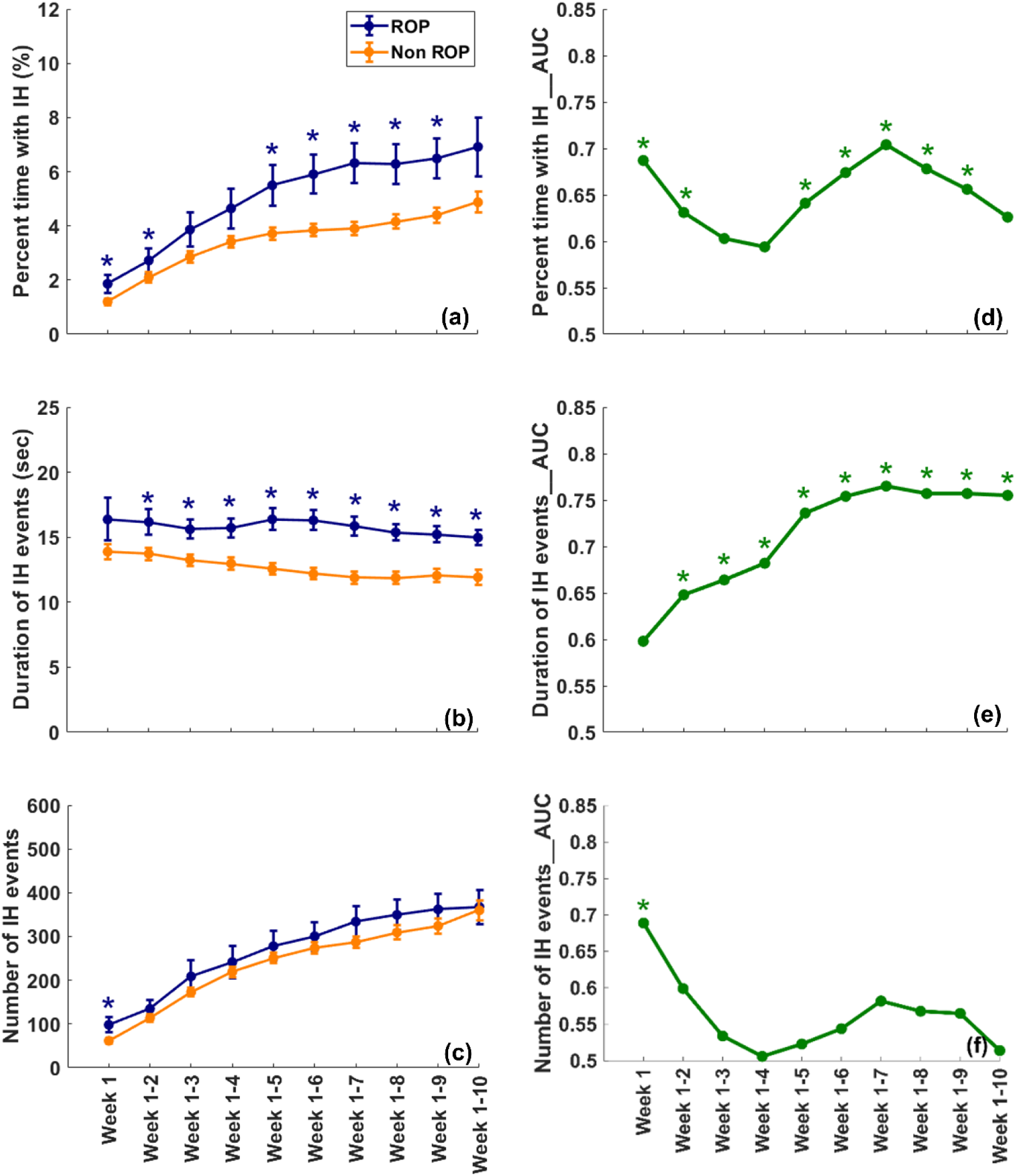
Cumulative process #2 results. (a)-(c): Cumulative comparison results (mean value ± SEM) between the ROP and non-Type1 ROP groups in IH variables, including the percent time with hypoxemia, duration of IH events, and number of IH events, respectively. * Indicates p < 0.05 for comparing the mean values between the two groups based on the Mann-Whitney U test. (d)-(f) Cumulative AUC results in IH variables, including the percent time with hypoxemia, duration of IH events, and number of IH events, respectively. * Indicates p < 0.05 for assessing whether a single IH variable is predictive (i.e., AUC > 0.5) based on a test which is equivalent to the Mann-Whitney U test.

